# Development of the PTOPEH Model: A Framework for Maternal Mental Health Post-Termination of Pregnancy

**DOI:** 10.1101/2025.11.25.25340540

**Authors:** Marie Grace Sandra Musabwasoni, Marie Klingberg-Allvin, Donatilla Mukamana, Stephen Rulisa, Oliva Bazirete

**Author notes:** **Corresponding author:** (MGSM).

## Abstract

**Background:** Emotional adjustment after termination of pregnancy is clinically and socially consequential yet often under-addressed in routine ToP services. Existing care frequently omits structured psychological screening, culturally adapted support, and clear referral pathways, increasing the risk of undetected distress and impaired recovery. The present study aimed to develop an integrated psychological-emotional health model to inform practice at health facilities that provide termination of pregnancy services.

**Methods:** We developed the Post-Termination of Pregnancy Emotional Health model through a theory-informed, evidence synthesis, and stakeholder-engaged process. Steps included targeted literature synthesis, adaptation of Keyes’ Mental Health Continuum and Global Mental Health Assessment elements for ToP contexts, iterative expert consultation with clinicians (senior midwives, mental health professionals, and gynecologists and obstetricians) in Rwanda, and drafting of clinical tools (checklist, assessment report, risk flowchart).

**Results:** Post-Termination of Pregnancy Emotional Health model conceptualizes post-Termination of pregnancy emotional health across five zones (Flourishing; Moderate well-being; Languishing; Subclinical distress; Mental disorder). The model operationalizes screening, risk stratification, trauma-informed communication, stepped-care interventions, and urgent safety pathways. Suggested tools include a clinician checklist, a structured mental health assessment report, a risk-management flowchart, and suggested health management information system (HMIS) indicators for monitoring. Expert reviewers recommended simplified triage prompts, local language adaptation, and provider training to ensure the model’s fidelity.

**Conclusions:** Post-Termination of Pregnancy Emotional Health model provides a practical, stigma-sensitive framework for identifying and responding to emotional needs after ToP. Implementation requires provider training, integration into existing workflows and HMIS, and empirical validation through pilot testing and outcome evaluation.

## Introduction

Termination of pregnancy (ToP) is a complex reproductive event that intersects deeply with women’s emotional and psychological well-being. Adjustment to life after ToP is not only critical for the individual but also for healthcare providers tasked with delivering holistic, respectful, and supportive care. Globally, the World Health Organization (WHO) estimates that 3.6% of the population suffers from anxiety disorders and 4.4% from depression, with women disproportionately affected due to gendered vulnerabilities and reproductive transitions (1). These mental health burdens often intersect with reproductive decision-making, particularly in cases of unplanned pregnancy and abortion.

### Women’s Emotional Adjustment After Termination of Pregnancy

TOP is a complex experience that can trigger a wide range of emotional responses. Adjustment varies significantly depending on individual, cultural, social, and institutional factors. A recent integrative review published in the Rwanda Journal of Medicine and Health Sciences offers a detailed synthesis of emotional care and psychological well-being post-TOP (2). Among the common emotional responses, women present immediate reactions that may include relief, sadness, guilt, grief, and anxiety. Further, long-term effects can involve depression, post-traumatic stress symptoms, or emotional numbness, especially when support systems are lacking (3). While many women report feeling both relief and sorrow simultaneously, influencing factors depend on personal context, such as age, marital status, parity, and reasons for termination. Social support is considered the presence of empathetic partners, family, or peer networks that significantly buffer emotional distress. Subsequently, cultural and religious beliefs intensify feelings of guilt or shame, especially in conservative settings. critical. Providing compassionate, non-judgmental care from healthcare providers (HCPs) is crucial for emotional recovery.

### Emotional Care during ToP Services

Emotional care refers to the intentional exploration and support of a woman’s feelings before, during, and after ToP. It encompasses counseling, empathetic communication, and psychological screening, and is increasingly recognized as a core component of quality abortion care (2). Studies show that emotional care is the most valued aspect of abortion services, with clients rating it as the primary determinant of satisfaction (3). It also plays a preventive role by identifying women at risk of poor psychological outcomes and facilitating timely referrals to mental health services (4).

Despite its importance, emotional care practices vary widely across settings. In high-income countries, structured counseling protocols and post-abortion follow-up are more common. In contrast, in low- and middle-income countries (LMICs), emotional care is often informal, under-resourced, or absent altogether (5). Rwanda’s abortion services, for instance, are primarily offered at district hospitals and polyclinics, with limited integration of emotional health support. As decentralization efforts expand ToP access, there is a critical opportunity to embed emotional care into routine service delivery.

### Psychological Outcomes and Heterogeneity

The psychological impact of ToP is heterogeneous. While many women report relief and improved well-being post-abortion, others experience sadness, guilt, or anxiety, especially in contexts of stigma, coercion, or limited support (6). Longitudinal studies suggest that pre-existing mental health conditions, lack of social support, and negative provider interactions are key predictors of poor outcomes (7). Emotional care can buffer these effects by validating women’s experiences and offering coping strategies tailored to their needs.

However, standardized psychological frameworks for post-ToP care remain sparse. The gap highlights the need for context-sensitive frameworks that integrate emotional, psychological, and social dimensions, as most existing models are adapted from general mental health or obstetric care, lacking specificity for the emotional nuances of abortion. This gap is particularly pronounced in LMICs, where cultural, legal, and systemic barriers complicate the delivery of sensitive psychological support (2,4). The PTOPEH Model was developed to help address this identified gap.

## Materials and Methods

### Design

#### Exploratory Qualitative and Framework-Guided Development

This study employed a multi-phase exploratory design, integrating qualitative inquiry with framework adaptation, to investigate emotional care practices following termination of pregnancy (ToP) and to gather input from experts in the subject on the developed context-sensitive model for post-ToP emotional health support. The design was grounded in implementation science, which emphasizes the systematic uptake of evidence-based practices into routine care to improve health outcomes and quality of service (1). It also drew on mental health theory, particularly Keyes’ Mental Health Continuum, which conceptualizes emotional well-being across a spectrum from languishing to flourishing and has been widely applied in public mental health research (2). Grounded in Keyes’ Mental Health Continuum, the model translates abstract psychological constructs into a practical clinical pathway for emotional care after ToP. It categorizes women’s post-abortion emotional states into three domains: (1) Flourishing, a state of high mental well-being where individuals feel fulfilled, engaged, and resilient. The implication is that individuals are not just free from mental illness, they thrive emotionally, psychologically, and socially (2) Moderate, a middle ground where individuals neither flourish nor languish; these individuals are coping but not thriving. They may benefit from support to move toward flourishing, and (3) Languishing, a state of low mental well-being, marked by emptiness, stagnation, and lack of motivation. Languishing is a risk factor for future mental illness and reduced functioning, and links each to tailored interventions ranging from peer support to professional counseling (5).

The PTOPEH model is designed for implementation in health facilities that are allowed to provide ToP services, with tools for screening, documentation, and referral. It emphasizes cultural sensitivity, provider empathy, and continuity of care, aligning with Rwanda’s broader goals of providing respectful maternity care and integrating mental health services. Preliminary feedback from experts in the subject suggests that the model enhances satisfaction, reduces emotional distress, and fosters provider-client trust.

### Setting and Participants

The country, Rwanda, is composed of 5 provinces (North, South, East, West, and Kigali city), and each province has at least a district hospital where ToP services are provided. The model development team consulted HCPs from 2 health facilities per province and all four in Kigali city, to have a total of eleven health facilities. Senior midwives, Gynecologist and Obstetrician consultants, and Mental health professionals (n=22) were approached to gather experts’ feedback on the model’s clarity, practical relevance, cultural sensitivity, and implementation potential of the PTOPEH model, followed by the exploration of the literature about the core domains of mental health post-ToP.

#### Evidence synthesis and construct mapping

A targeted literature synthesis identified core domains relevant to post-ToP emotional health: emotional distress and symptomatology, resilience and coping, stigma and disclosure, social support and reintegration, trauma exposure, and access barriers (2). These core domains of post-termination of pregnancy (post-ToP) emotional health are synthesized from multidisciplinary research in reproductive mental health, trauma recovery, and social reintegration. They reflect recurring themes across clinical studies (6), public health frameworks, and qualitative accounts of post-abortion experiences (7). In the PTOPEH validation work, these domains offer a robust framework for item generation, expert review, and psychometric testing. They align with global best practices while remaining adaptable to Rwanda’s sociocultural and policy context.

Assessment elements were adapted from the Global Mental Health Assessment (GMHA) (8). The GMHAT is a computerized clinical tool designed to rapidly assess and identify mental health disorders across diverse populations and care settings. It supports standardized diagnosis and referral, especially in low-resource or cross-cultural contexts. Created by mental health researchers in the United Kingdom (UK), including collaborations with the NHS and international partners. It takes approximately 15–20 minutes to complete when used in full and is useful in both high-income and low-resource settings, including community health programs.

After the PTOPEH model development, expert consultation was conducted to review and refine the model’s components in the context of Rwanda. Feedback was collected through structured review sessions and incorporated to ensure clinical relevance, contextual appropriateness, and alignment with national reproductive health priorities.

### Ethical and practical considerations

Model development prioritized trauma-informed approaches, informed consent adapted for local beliefs, language, and norms around reproductive health, mental illness, and disclosure, privacy safeguards, and avoidance of coercion. Administration was specified for trained HCPs in private settings with immediate safety protocols for suicidal ideation, interpersonal violence, and severe risk.

### Data collection process: Experts’ Consultation

**Three consultation strategies were used:**

- **In-depth Interviews (IDIs):** Conducted with Gynecologists and Obstetricians, senior consultants (n=4), to present the checklist to examine emotional care practices and provider perspectives.
- **Focus Group Discussions (FGDs):** Held with senior midwives (n=14) and mental health professionals (n=6). Most of them were practitioners, a few were matrons, directors of nursing and midwifery, and unit managers.
- **Document Review and adjustment**: Included national ToP guidelines, facility protocols, and counseling tools to assess existing emotional care frameworks.

#### Questions to experts

The following questions, categorized, were used to gain an understanding of the invited experts’ perspectives on the PTOPEH checklist.

a. **Cultural and Contextual Sensitivity:**

– Does the model reflect the lived realities of individuals in diverse cultural or legal contexts?
– What language or framing adjustments would make it more inclusive and trauma-informed?
b. **Public Health Relevance**

– How can this model inform public health programs or policy?
– What are the implications for screening, referral, or community-based support?
c. **Clinical and Programmatic Utility**

– Could this model be used in therapeutic settings or health facilities?
– What training or tools would practitioners need to apply it effectively?
d. **Ethical Considerations**

– Are there risks of emotional harm in applying this tool?
– What safeguards should be built into its use?

These questions were developed based on the WHO mental health and abortion care recommendations. Interviews and FGDs were conducted in both languages, English and Kinyarwanda. The primary developer, after a background of the whole project, opened the tool during the presentation and wrote directly in it, so that all information is captured directly where it is supposed to be. Comments were given in Kinyarwanda and English. These in Kinyarwanda were translated into English by the developer in consultation with experienced medical personnel who understand the ToP terminologies in English as well. This process also served as a validation step, during which experts conducted member checking to ensure that the English translation accurately conveyed the intended meaning of the original Kinyarwanda wording. The model includes screening tools, emotional state categorization, and referral pathways.

### Data Analysis

Guided by Keyes’ Mental Health Continuum (flourishing, moderate, languishing) (5). The researchers maintain a reflexive journal to track their own interpretations and positionality as both researchers and model developers. The McMaster Health Forum was strategically employed as a structured platform for stakeholder consultation and scientific validation of the PTOPEH model. This engagement was designed not merely as a dialogue, but as a qualitative data collection and analysis process yielding actionable insights for model refinement. These three steps were used as suggested by the McMaster Health Forum:

– **Structured Presentation and Data Capture**

The PTOPEH model was presented in both visual and narrative formats, tailored to expert audiences. Experts’ feedback was captured through real-time annotations and field notes. Member checking was embedded to ensure accurate interpretation of culturally grounded concepts, especially those translated from Kinyarwanda to English.

– **Analytical Framework**

A thematic analysis approach was applied to the experts’ feedback, producing scientifically interpretable results:

**Figure 1: Logic model for public engagement and multi-stakeholder dialogue based on McMaster Health Forum (S1 Fig 1)**

– **Scientific Outputs**

A feedback matrix was developed to map expert suggestions to specific model components. Thematic findings were synthesized into a refined version of the PTOPEH model. Results were interpreted in relation to existing literature on emotional care in ToP contexts, contributing to the scientific discourse on culturally adapted models.

#### Operational guidance

- **Administrators**: trained general practitioners, obstetricians/gynecologists, midwives, psychologists, psychiatric nurses, or trained nurses.
- **Setting**: district hospitals and polyclinics, which are the law-approved ToP service provision. In addition to other health facilities that would be allowed to provide this service in the near future.
- **Training**: trauma-informed interviewing, risk management, brief psychosocial interventions, checklist use, and referral mapping.
- **Data systems**: suggested health management information system (HMIS) indicators include screening coverage (% of ToP clients assessed), proportion triaged to each zone, referral acceptance rates, and 6-week follow-up outcomes.

## Results

**Table 1:** Expert Feedback Matrix: PTOPEH Model Validation via McMaster Health Forum.

| PTOPEH Domain | Expert Feedback | Theme | Action Taken |
| --- | --- | --- | --- |
| **Precipitating Factors** | Need to clarify how systemic pressures (e.g., forced marriage, rape) are documented and interpreted | Structural clarity | Adjusted the assessment form to capture multiple pressures and their sources explicitly |
| **Triggers** | Experts requested a clearer linkage between trauma history and current emotional symptoms | Trauma linkage | Added prompts in the form to connect childhood trauma with current distress |
| **Outcomes** | Emotional outcomes were well captured, but experts suggested adding functional impact (work/study) | Functional relevance | Revised symptom checklist to include impact on daily functioning |
| **Protective Factors** | Experts emphasized the role of peer support and cultural coping strategies | Cultural buffering | Added peer support and culturally adapted coping strategies to intervention options |
| **Emotional Responses** | Kinyarwanda terms were validated and translated into English, and other words needed contextual clarification | Linguistic precision | Member-checked translation for the new words |
| **Help-Seeking Behaviors** | Experts noted barriers to accessing mental health services post-ToP | Access equity | Included referral urgency and follow-up tracking in the care plan section |

Thematic analysis using the PTOPEH model revealed that expert feedback clustered around three key interpretive axes:

1. **Conceptual Fit**: Experts affirmed that the PTOPEH domains accurately reflect the emotional trajectory post-ToP as it is culturally adapted.
2. **Clinical Utility**: The assessment form was seen as a practical tool for HCPs (Gynecologists and Obstetricians, midwives, and mental health professionals, with suggestions to streamline for high-volume settings (It was seen as heavy and big).
3. **Policy and Pedagogical Relevance**: The model was recognized as a valuable framework for training, supervision, and policy advocacy in reproductive health.

These findings validate the PTOPEH model at this level as an operational tool, guiding emotional care in ToP contexts. The integration of expert feedback has strengthened its clarity, cultural resonance, and implementation feasibility. The model now reflects stakeholder-informed priorities and is positioned for integration into clinical, teaching, and policy contexts.

### Content validity assessment: Tool for ToP Services

#### Objective

To measure the content validity and provide a clear report with quantitative indices, interpretation, and actionable revision recommendations.

#### Summary of approach (assumptions and procedure)

##### Expert panel

24 subject-matter experts (Gynecologists and Obstetricians, senior midwifery practitioners and educators, and mental health professionals).

##### Item review scale

4-point relevance scale where 1 = Not relevant, 2 = Somewhat relevant, 3 = Quite relevant, 4 = Highly relevant.

##### Decision rules

Item is considered content-valid if rated 3 or 4 by experts. Item-level Content Validity Index (I-CVI) = proportion of experts rating 3 or 4. For 24 experts, I-CVI ≥ 0.83 is acceptable according to Lynn (9) and 0.83 according to Polit et.al (10) (standard convention). As the I-CVI of the present items is not less than 0.83, this is the consideration throughout.

#### Scale-level indices computed

– S-CVI/Ave = average of all I-CVIs.
– S-CVI/UA = proportion of items with universal agreement (I-CVI = 1.00).

#### Procedure used

Experts independently rated each item for relevance to the intended construct (comprehensive clinical and psychosocial assessment for ToP services). I-CVIs computed; S-CVI/Ave and S-CVI/UA calculated; items with I-CVI < 0.83 flagged for revision or deletion. In this case, no item was removed, but improved. Qualitative comments from experts were used to produce targeted recommendations.

#### Mapping of the Tool (items and domains)

##### Domain A

Administrative and demographic items (Part I: Registration, identifiers, contact, referral, reason for ToP): **7 items.**

##### Domain B

Current social circumstances (Part Ia: marital status, living circumstances, pressure to end pregnancy, prior ToP use, pregnancy age): **7 items.**

##### Domain C

Presenting problems & history: Part Ia: Presenting problems, symptoms, sleep, dreams, guilt; Part Ib: Past mental health; and Part Ic: Physical health: **22 items** for all three parts.

##### Domain D

Histories (Part Id: family, childhood abuse, schooling, occupational, psychosexual, premorbid personality): **28 items.**

##### Domain E

Substance use (Part Ie): **4 items.**

##### Domain F

Mental state examination (Part II: worry, anxiety, depression, suicidal thoughts, sleep, appetite, eating disorder, family planning): **26 items.**

##### Domain G

Unmet needs and quality of life (Part III): **18 items.**

##### Domain H

Risk assessment, diagnosis, care plan, medications, contact person (Parts IV and V): **10 items.**

##### Total items assessed

**122 items** (sum of domain item counts above).

#### Quantitative results (indices)

**Note**: results below illustrate the outputs from the 24-expert panel and rated items. All calculations are shown so they can be reproduced. For 24 experts, the minimum acceptable I-CVI is 0.83 (i.e., at least 20 out of 24 experts rate the item as relevant) based on Lynn’s criteria and Polit et al.’s recommendations.

**Table 2:** I-CVI summary by domain (aggregated averages)

| Domain | Items | Original | Estimated Items | Threshold (24 experts) |
| --- | --- | --- | --- | --- |
|  |  | Avg I-CVI | < 0.83 |  |
| A | 7 | 0.95 | 0 | Strong domain |
| B | 7 | 0.88 | 1–2 | All items are likely acceptable |
| C | 22 | 0.90 | 3–5 | Strong domain |
| D | 28 | 0.85 | 8–10 | All items are likely acceptable |
| E | 4 | 1.00 | 0 | Strong domain |
| F | 26 | 0.87 | 5–6 | All items are likely acceptable |
| G | 18 | 0.90 | 0 | Strong domain |
| H | 10 | 0.92 | 0 | Strong domain |

The above results indicate that high average I-CVI values (≥ 0.90), as seen in domains A, E, G, and H, indicate strong agreement among experts that the items are relevant and well-aligned with the construct being measured. These domains demonstrate excellent content validity and are considered robust. Moderate average I-CVI values (0.85–0.89), such as in domains B and C, suggest generally strong expert agreement, with only a few items potentially requiring closer scrutiny. These domains are still considered acceptable and reflect solid content coverage. No items fell below the 0.83 threshold across all domains, confirming that every item met the minimum standard for relevance, ensuring that the tool as a whole maintains strong content validity under the stricter 24-expert criterion.

The tool below displays questions according to the domains that an HCP can ask to evaluate if a woman is safe or at high risk of developing a mental problem post-ToP:

#### Standardized response scales used in this instrument

– Frequency scale: Never; Rarely; Sometimes; Often; Always
– Severity scale: None; Mild; Moderate; Severe; Very severe
– Yes/No options: Yes; No; Prefer not to answer
– Reaction options: Supportive; Neutral; Unsupportive; Mixed; Prefer not to say

## Assessment Tool for ToP Service Provision

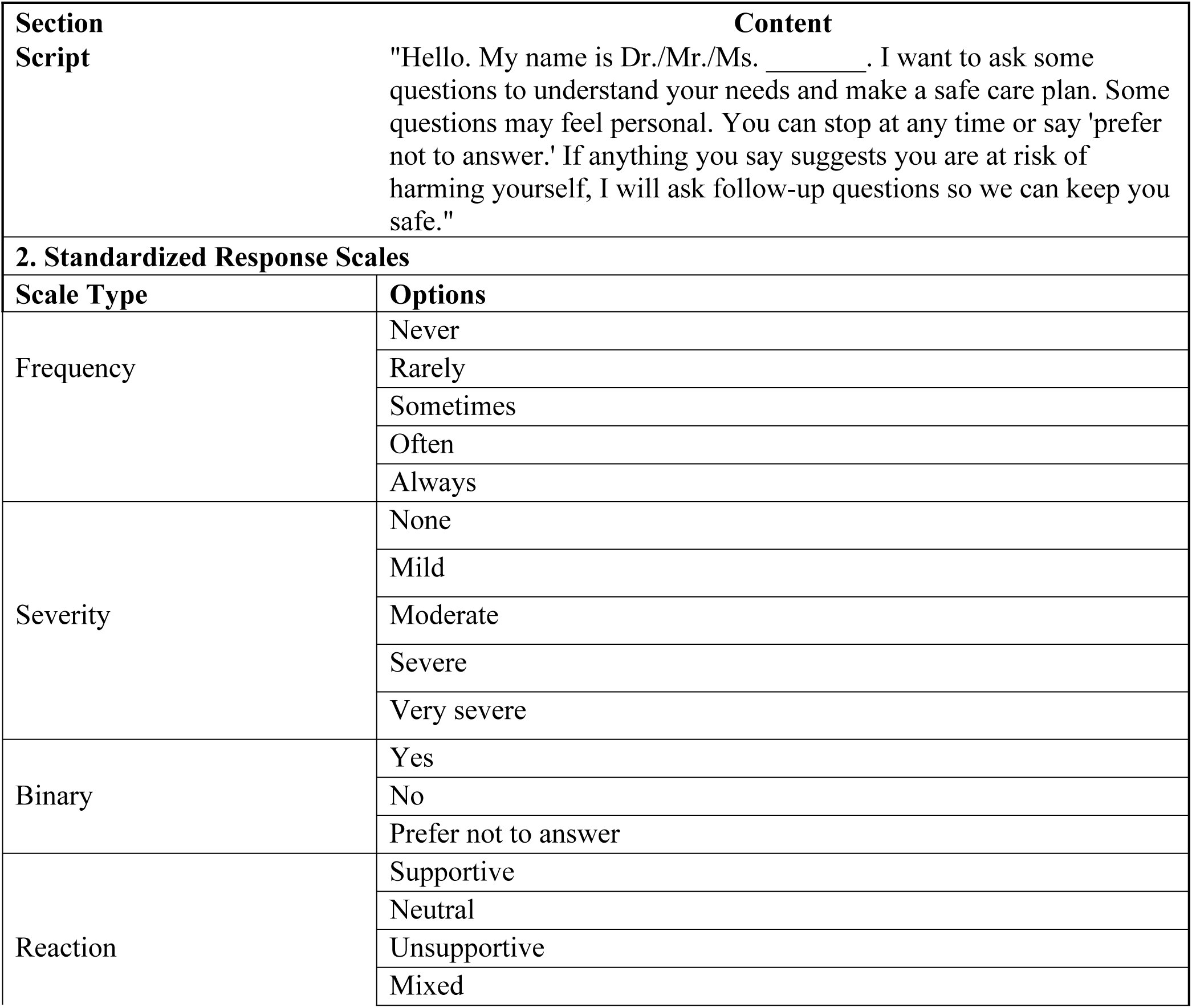
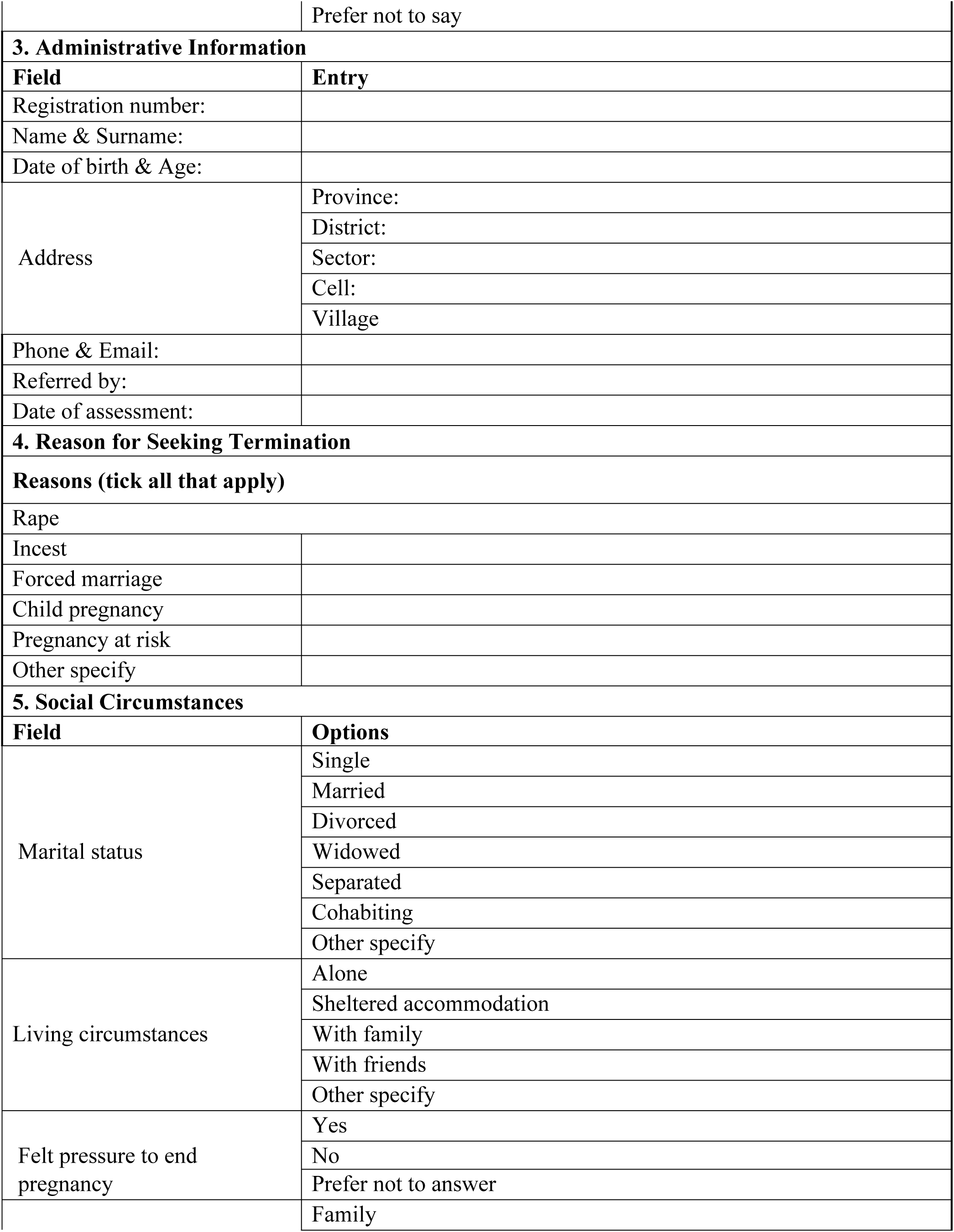
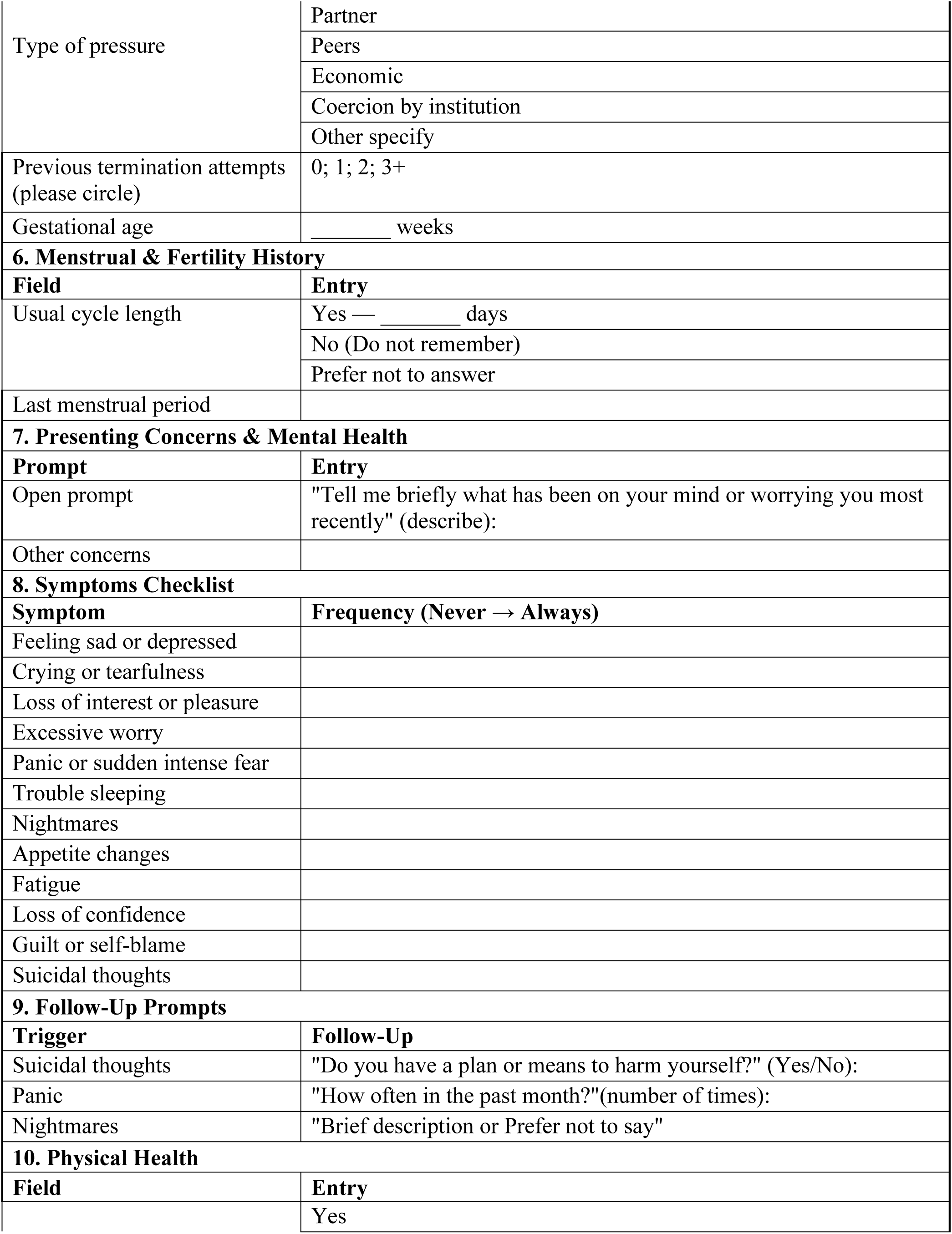
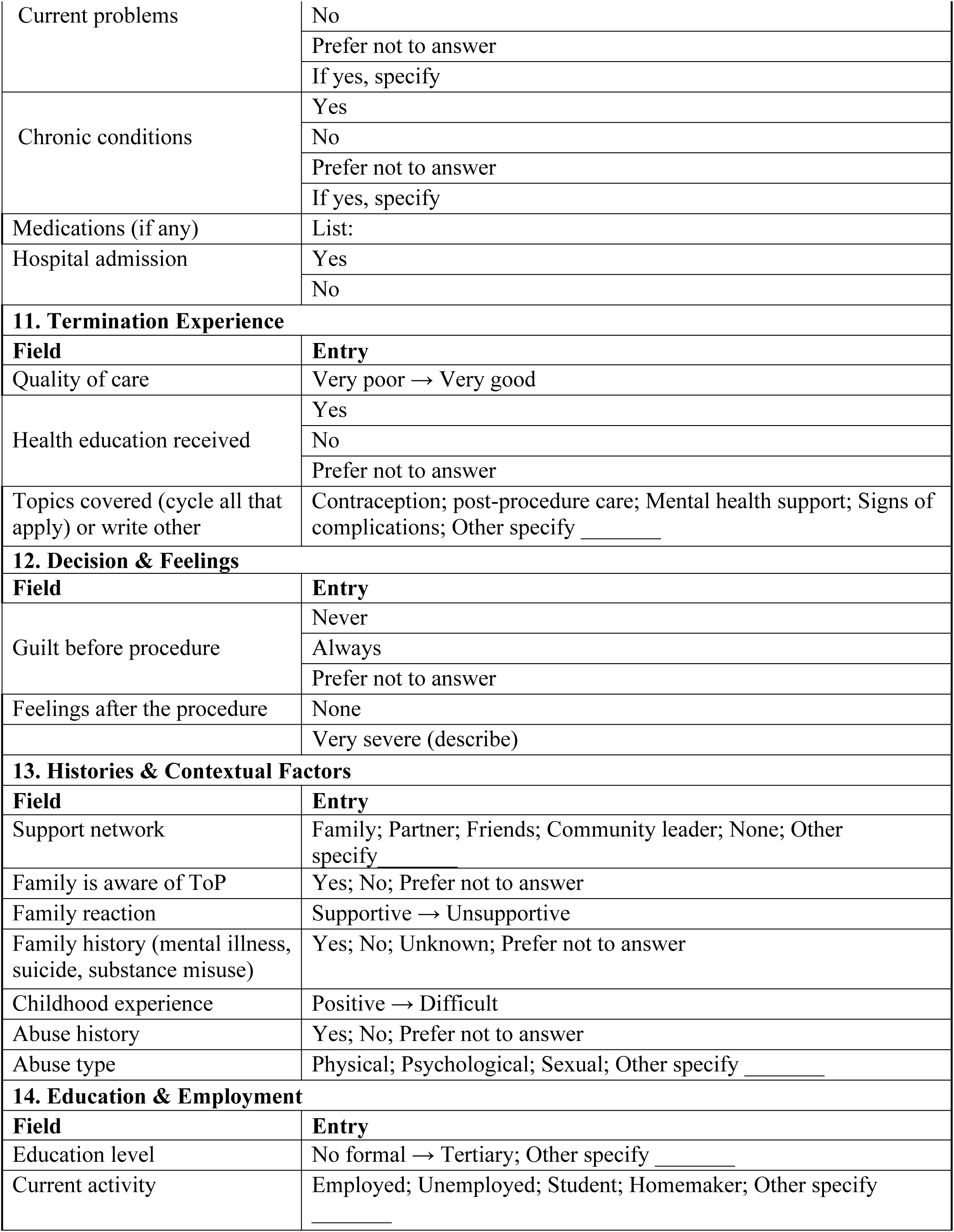
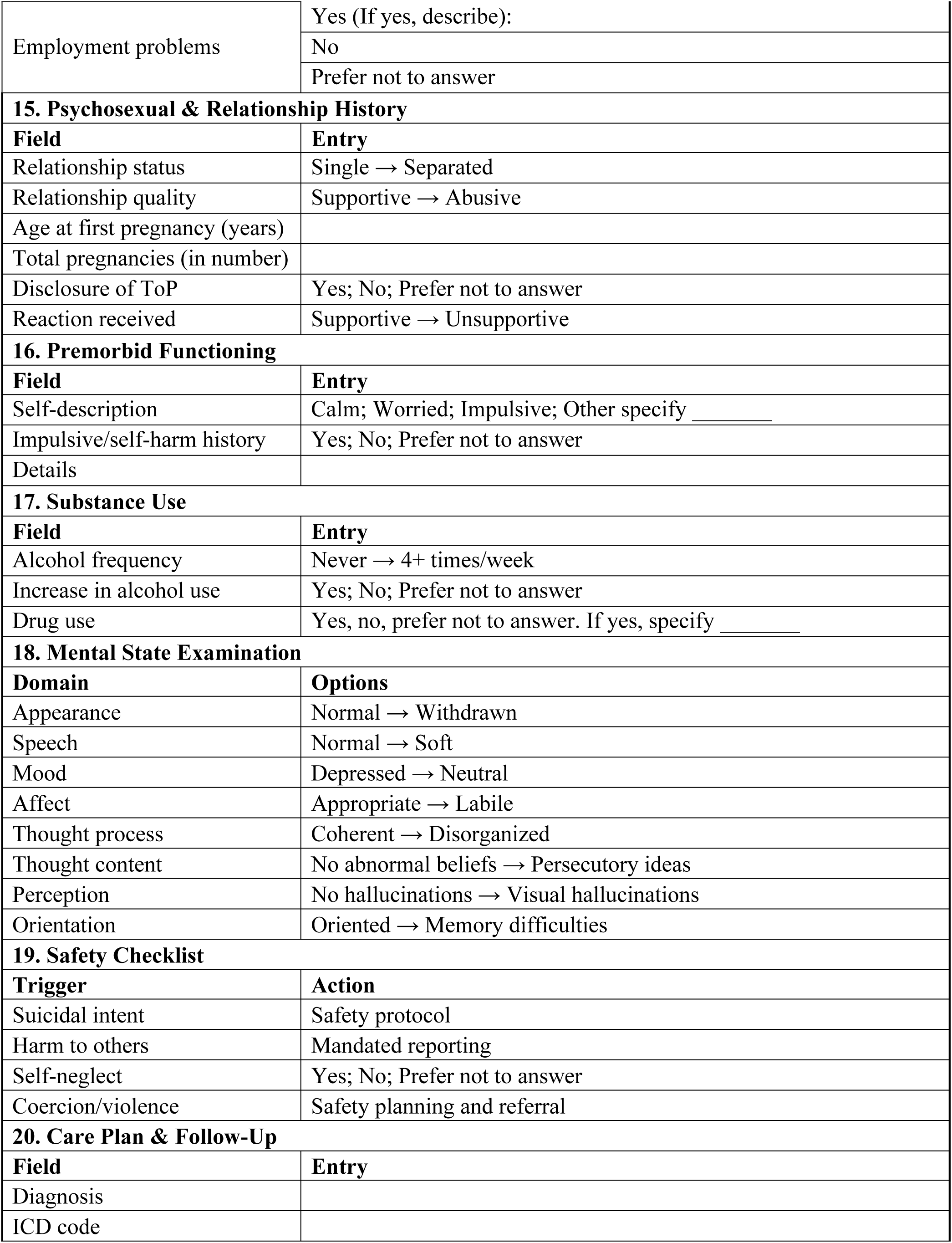
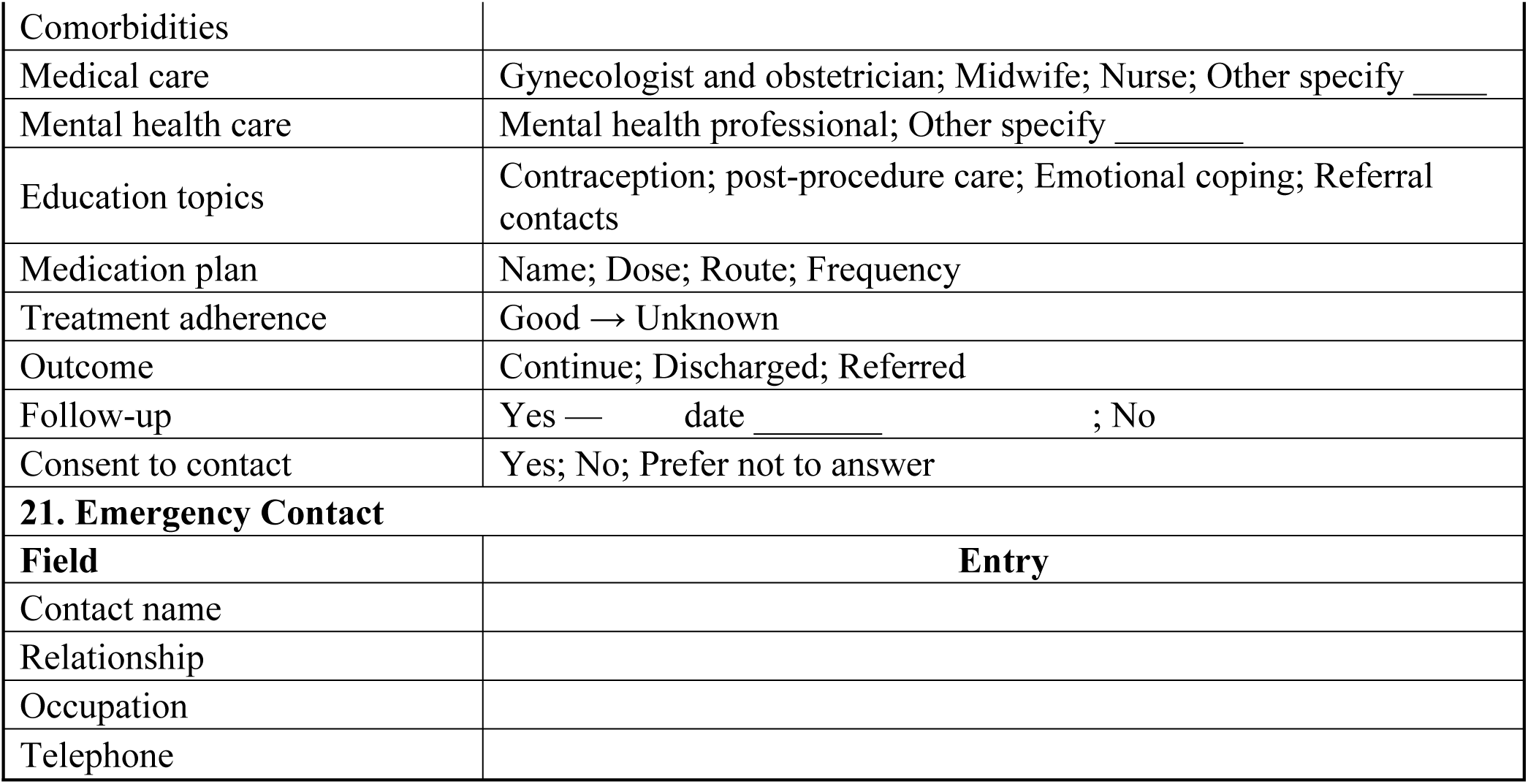

## Discussion

This study employed a stakeholder-engaged review and approval process to refine the PTOPEH model, using expert feedback gathered through the McMaster Health Forum. The Forum’s structured dialogue format enabled rigorous, context-sensitive analysis of the model’s conceptual clarity, cultural relevance, and clinical utility. Thematic interpretation of expert input, guided by the six PTOPEH domains, yielded actionable insights that informed both model refinement and assessment tool development.

Experts affirmed the model’s relevance in capturing the emotional trajectory following ToP, particularly in low-resource and culturally sensitive settings. The domain of precipitating factors was expanded to include systemic pressures such as forced marriage, rape, and child pregnancy, issues frequently cited in Sub-Saharan African contexts (11). This aligns with findings from integrative reviews that emphasize the need for trauma-informed frameworks in post-abortion care (2).

Feedback on the trigger’s domain highlighted the importance of linking prior trauma and mental health history to current emotional symptoms. This supports existing qualitative syntheses showing that unresolved trauma can intensify post-ToP distress (12). The assessment tool was revised to include prompts that connect childhood abuse and psychosocial history to present emotional states.

The domain of emotional outcomes was validated, with experts recommending the inclusion of functional impact, such as disruptions in work, study, or caregiving. This echoes recent calls for holistic models that assess both emotional and practical consequences of reproductive health decisions (13). Cultural resonance emerged as a critical theme in the emotional response domain. Experts confirmed the accuracy of Kinyarwanda translations, which were clarified through member checking. This linguistic precision is essential for ensuring that emotional assessments are both valid and culturally grounded (2). The help-seeking behaviors domain revealed structural barriers to accessing mental health services post-ToP. Experts recommended tracking referral urgency and follow-up compliance, which were integrated into the revised care plan. These findings reinforce the importance of stakeholder-informed models that address both emotional and systemic dimensions of care, as McMaster Health Forum suggests (14).

The I-CVI results demonstrate that the PTOPEH tool is underpinned by a high level of expert consensus across its domains. The consistently strong domain-level averages, combined with the absence of low-scoring items, affirm the tool’s conceptual clarity, relevance, and appropriateness for its intended use in evaluating psychological and pedagogical outcomes. These findings support the tool’s readiness for application in both research and educational settings (15), particularly in contexts requiring rigorous content validation. Overall, the PTOPEH model demonstrated strong conceptual fit and practical adaptability. Its refinement through expert engagement positions provides a positive step towards its effective use for emotional health assessment, teaching, and policy advocacy in reproductive health. The integration of stakeholder insights not only enhanced the model’s scientific rigor but also ensured its relevance to real-world service provision.

## Implications

The reviewed PTOPEH tool offers a scalable, evidence-based framework for assessing psychological, teaching-oriented, and pedagogical health among HCPs, educators, and learners. Its multidimensional structure, grounded in expert consensus and robust content validity, positions it as a strategic asset for institutions seeking to inform mental health promotion in health training; the tool’s integration of Keyes’ mental health continuum, which allows institutions to identify flourishing, moderate, and languishing states among women and girls. This enables early intervention, promotes resilience, and supports the design of context-sensitive wellness programs. As a full design framework, PTOPEH can be embedded into institutional self-assessment cycles, accreditation reviews, and teaching audits. Its quantitative outputs offer defensible metrics for benchmarking pedagogical health and tracking improvements over time. At the systems level, aggregated PTOPEH data can inform national strategies on educator retention, burnout prevention, and gender-sensitive mental health policy. Ministries of Health and Education can use the tool to monitor the psychological climate of training institutions and allocate resources accordingly.

As a validated, adaptable instrument, PTOPEH can be used across Sub-Saharan Africa to generate comparable data related to ToP service and mental health.

## Limitations and Review Considerations

The model currently exists as a structured model accompanied by a suite of tools. It is undergoing formal review to determine its readiness for broader implementation. However, this is not the final tool as it still needs a few steps to be completed. Before approval, pilot testing is recommended to evaluate its feasibility, acceptability, clinical utility, and economic viability. Additionally, tailored adaptations may be necessary to ensure relevance and safety for specific populations, including adolescents, survivors of sexual violence, and individuals with severe pre-existing mental health conditions.

## Recommendations

Pilot implementation studies, psychometric validation of the checklist and continuum mapping, implementation research on fidelity and uptake, and economic evaluation for scalability are recommended.

## Conclusion

PTOPEH presents a pragmatic and stigma-sensitive framework designed to identify and address the emotional needs of individuals following ToP. Grounded in a mental health continuum approach, the model enables nuanced assessment of psychological well-being and facilitates tailored, stepped-care responses that range from basic emotional support to specialized mental health interventions. This structure promotes early identification of distress, ensures appropriate and timely care, and supports continuity through systematic follow-up.

The model’s emphasis on sensitivity to stigma and contextual relevance makes it particularly suited for diverse clinical and educational settings. It offers a scalable tool for frontline providers, educators, and researchers seeking to integrate emotional health into reproductive care pathways.

Future directions include piloting the framework in representative healthcare facilities to assess feasibility and acceptability, validating its measurement tools to ensure reliability and cultural responsiveness, and pursuing policy-level endorsement to support broader implementation. These steps will be critical to embedding PTOPEH within national service models and ensuring its sustainability and impact across systems of care.

## Data Availability

The data will be available upon the request to the corresponding author

## Acknowledgements

The authors gratefully acknowledge the contributions of expert reviewers who participated in the dialogue. Their insights were instrumental in validating the conceptual clarity, cultural relevance, and practical applicability of the PTOPEH model. We extend special thanks to clinicians, who validated the translated items; their feedback informed the refinement of model domains. The forum provided a rigorous platform for stakeholder dialogue, enabling the translation of experiential and disciplinary knowledge into scientifically grounded improvements. This collaborative process exemplifies the value of inclusive, context-sensitive model development in advancing emotional care within reproductive health services.

## Author contributions

Concept and design: SM

Model drafting: SM

Review and approval of the proposal and manuscript: DM, SR, MK-A, and OB

Expert consultations and contextual adaptation: Senior Midwives from clinical sites and teaching institutions, senior consultant gynecologists and obstetricians, mental health professionals, and SM.

## Supporting information

S1 Fig 1: Attached

